# Synthesized Economic Evidence on the Cost-effectiveness of Screening Familial Hypercholesterolemia: A meta-analytic review and Aggregate Distributional Cost-Effectiveness Analysis

**DOI:** 10.1101/2023.12.09.23299771

**Authors:** Mengying Wang, Shan Jiang, Jiao Lu, Kai Tan, Yuanyuan Gu, Shunping Li

## Abstract

**Background:** Familial hypercholesterolemia (FH) is a prevalent genetic disorder with global implications for severe cardiovascular diseases. Amid ongoing advancements in genetic disease screening and treatment, the economic evaluation of FH is increasingly vital. Despite numerous studies, international disparities persist, necessitating a comprehensive analysis of the economic assessments of FH screening to provide valuable insights. This study aims to globally examine economic assessments of FH, synthesize evidence, and present economic insights into the impact of FH screening on population equity.

**Methods:** Systematic literature analysis was used to systematically evaluate 19 FH screening economic evaluation studies and synthesize the evidence. Meta-analysis was used to comprehensively explore the total cost-effectiveness and total net health benefit of different FH screening methods. The aggregate Distributional Cost-Effectiveness Analysis (DCEA) was utilized to assess the impact of FH screening on population equity.

**Results:** The study results reveal significant differences in the economic evaluations of FH across different countries, and we provide detailed descriptions of unique characteristics. The comprehensive results of cost-effectiveness analysis indicate that FH screening is cost-effective in the majority of countries. The meta-analysis synthesizes the economic impacts of Cascade Screening and Universal Screening across various health outcomes. Additionally, our aggregate Distributional Cost-Effectiveness Analysis (DCEA) suggests that FH screening strategies have the potential to alleviate health inequalities.

**Conclusion:** This study provides a global perspective on FH economic evaluation. Despite international differences, findings suggest that, in the majority of cases, FH screening is cost-effective, improving health equity and overall population health. The study offers a positive outlook for future health decisions.

## INTRODUCTION

Familial hypercholesterolemia (FH) is a grave autosomal dominant inherited condition marked by elevated plasma concentrations of low-density lipoprotein (LDL) cholesterol.^1^ The pathogenesis of FH arises from mutations in the genes encoding for the low-density lipoprotein receptor (LDLR), apolipoprotein B (APOB), and proprotein convertase subtilisin/kexin type 9 (PCSK9).^2,3^ These genetic anomalies disrupt cholesterol metabolism within patients, leading to significantly raised plasma LDL cholesterol concentrations.^4^ As a consequence, individuals with FH are predisposed to accelerated atherosclerotic cardiovascular disease (ASCVD) and coronary artery disease (CAD) development, which invariably augments their risk of premature mortality.^5^

Classifications of FH differentiate between homozygous familial hypercholesterolemia (HoFH) and heterozygous familial hypercholesterolemia (HeFH).^6^ Cumulatively, their global prevalence was historically estimated to be 1 in 500. However, more recent investigations suggest a prevalence nearing 1 in 200, equating to a total of 32 million FH patients or variant carriers worldwide.^7^ Alarmingly, despite its status as a prevalent genetic disorder, the majority of FH cases remain undiagnosed;^8^ in many countries, less than 1% of potential FH cases have been accurately identified.^9^

On an optimistic note, even though FH can have severe health implications, early detection coupled with prompt pharmacological interventions - such as with statins, ezetimibe, PCSK9 inhibitors, and the newly introduced bempedoic acid - can substantially reduce the risk of myocardial infarction in FH patients by up to 76%.^10,11^ Such interventions also thwart the precocious onset of atherosclerosis, thereby enabling these FH patients to lead lives with normal life expectancy.^12^ Hence, screening for FH is pivotal for preventing CVD events, enhancing life expectancy, and improving the overall quality of life in affected individuals.^7^

Some national health authorities and professional medical organizations are placing increasing emphasis on FH screening, endorsing actions through the formulation of guidelines and expert consensuses. For instance, in 2008, the National Institute for Health and Clinical Excellence (NICE) unveiled a UK guideline dedicated to the identification and management of patients with FH.^13^ Similarly, the European Atherosclerosis Society’s FH Studies Collaboration has recently underlined the imperative nature of a global registry for FH, advocating for concerted global initiatives.^14^ The Australasia Network Consensus Group, aiming to better guide clinicians in the management of FH, has also instituted novel directives.^15^ Northern Ireland, Scotland, and Wales have pioneered national FH services.^16^ While the importance of FH screening has garnered recognition among numerous professionals, there remains a significant gap concerning its economic implications. Notably, there is a dearth of comprehensive research assessing whether FH screening warrants inclusion in health insurance schemes.

Health inequality, as a pivotal policy concern within the global healthcare system, compels us to delve into the distribution of health costs and outcomes among diverse populations while evaluating the overall health input benefits of healthcare measures.^17,18^ Despite persistent calls for health technology assessment (HTA) agencies to incorporate fairness evaluations into their decision-making processes,^19^ there remains a lack of systematic research on the impact of FH screening on health equity. To address this knowledge gap, our study employs, for the first time, an aggregate Distributional Cost-Effectiveness Analysis (DCEA) methodology. This approach, introducing considerations of fairness atop traditional health economic evaluations, comprehensively explore the impact of FH screening strategies on health equity.^20^

Considering the knowledge gap, the overarching objective of this manuscript is to synthesize the economic evidence regarding (1) the cost-effectiveness of FH screening through meta-analytic review and (2) the impact on the equity across the population.

## METHODS

We employed a systematic review and data extraction approach to provide robust evidence for economic evaluation of FH screening strategies. Following this, a meticulous meta-analysis was undertaken to synthesize the gathered data. Subsequently, we applied an aggregate distributional cost-effectiveness analysis. This comprehensive analysis not only furnishes policymakers with detailed information but also quantitatively assesses the impact of health policies on health equity.

### 1. Systematic Review

#### Search strategy

In this study, we employed a rigorous literature search method to ensure the systematic retrieval and analysis of economic evaluation literature on FH screening. We used key terms and corresponding MeSH terms such as “familial hypercholesterolemia,” “cost-effectiveness analysis,” “disease screening,” and “health economics” to search multiple important databases, including PubMed, Web of Science, and Embase. The search cutoff date was October 20, 2023, and we limited the publication year of the literature to 2000 and beyond.

To ensure the accuracy and consistency of the research, we established predefined inclusion and exclusion criteria. Our inclusion criteria were as follows: (1) study participants being patients with FH, (2) the study involving FH disease screening, (3) the study conducting an economic evaluation of the screening methods, and (4) language restriction to English. Exclusion criteria included: (1) studies involving participants with multiple diseases simultaneously, (2) studies where the primary focus of the economic evaluation is not disease screening, (3) studies not reporting the process or results of economic evaluations, and (4) reviews, conference papers, disease guidelines, etc.

#### Data Extraction

Considering the diverse nature of the study literature, we utilized Excel tools for the meticulous extraction of data. The extracted information encompassed a wide array of variables, including background, perspective, currency unit, screening strategies, screening targets, treatment drugs, economic evaluation methods, cost-effectiveness analysis results, and intricate details regarding the decision models employed. The systematic extraction and integration of this comprehensive dataset were pivotal in furnishing comparable information, thereby augmenting the evidentiary foundation for our study (Table 1 and 2).

**Table 1.**
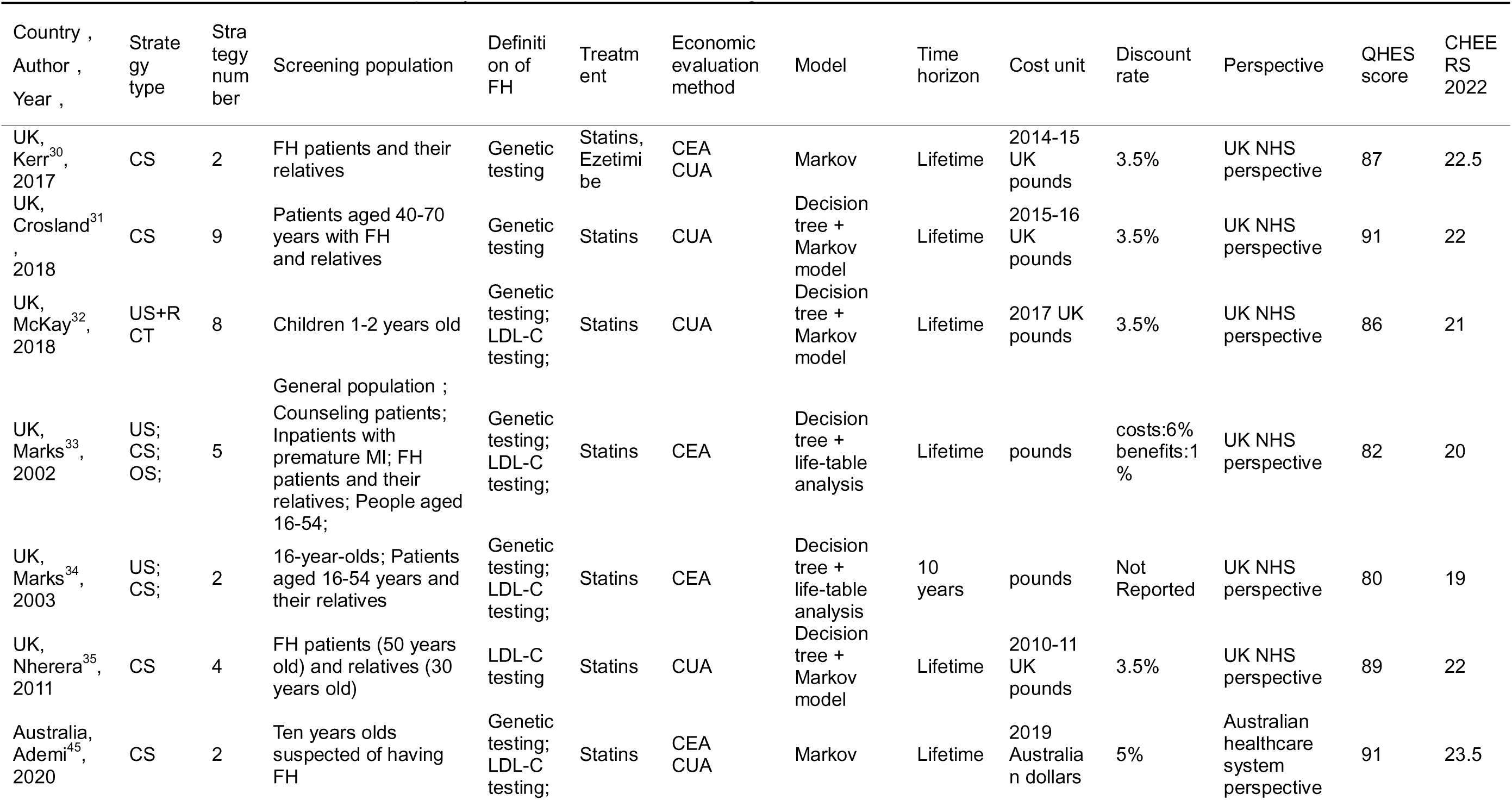
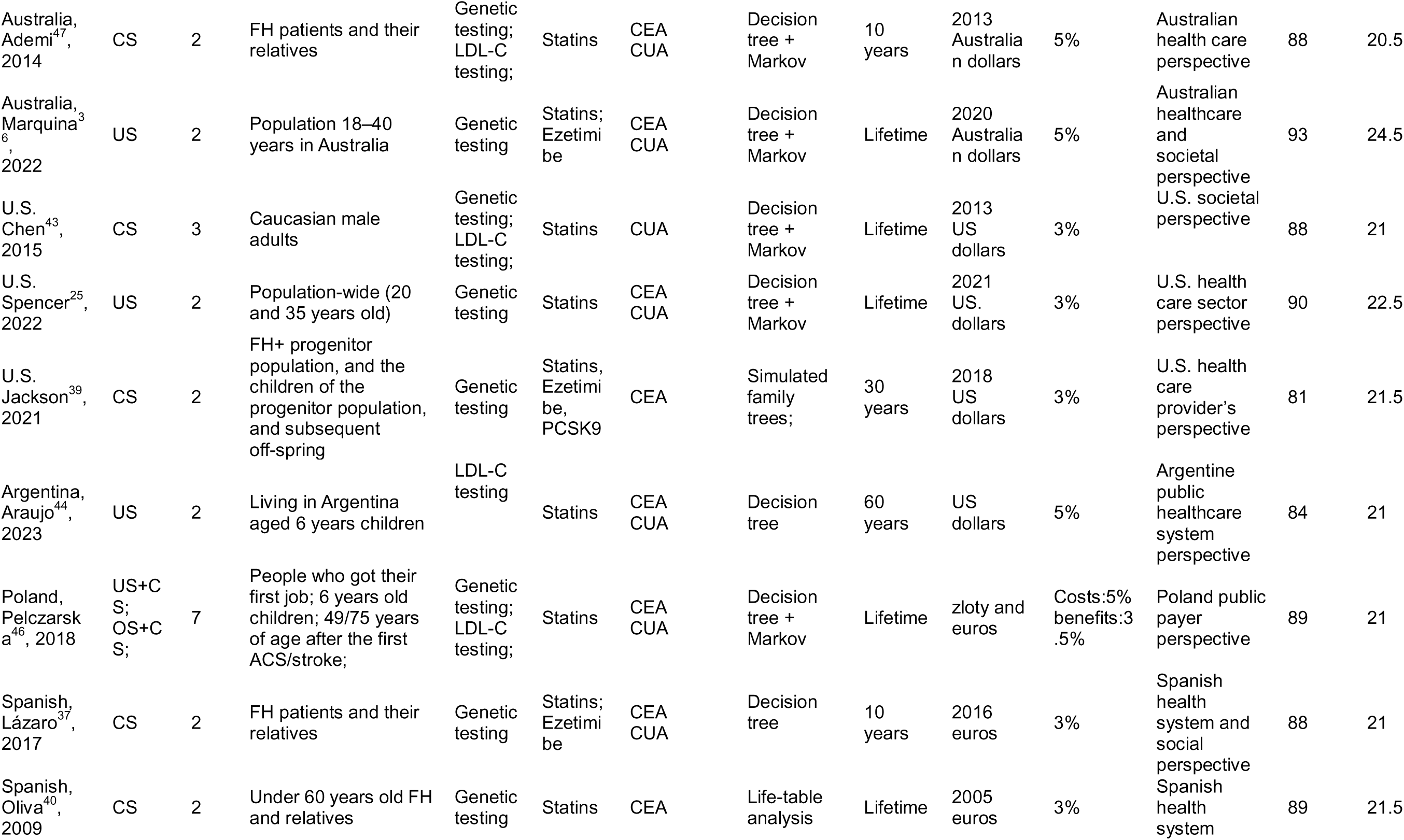
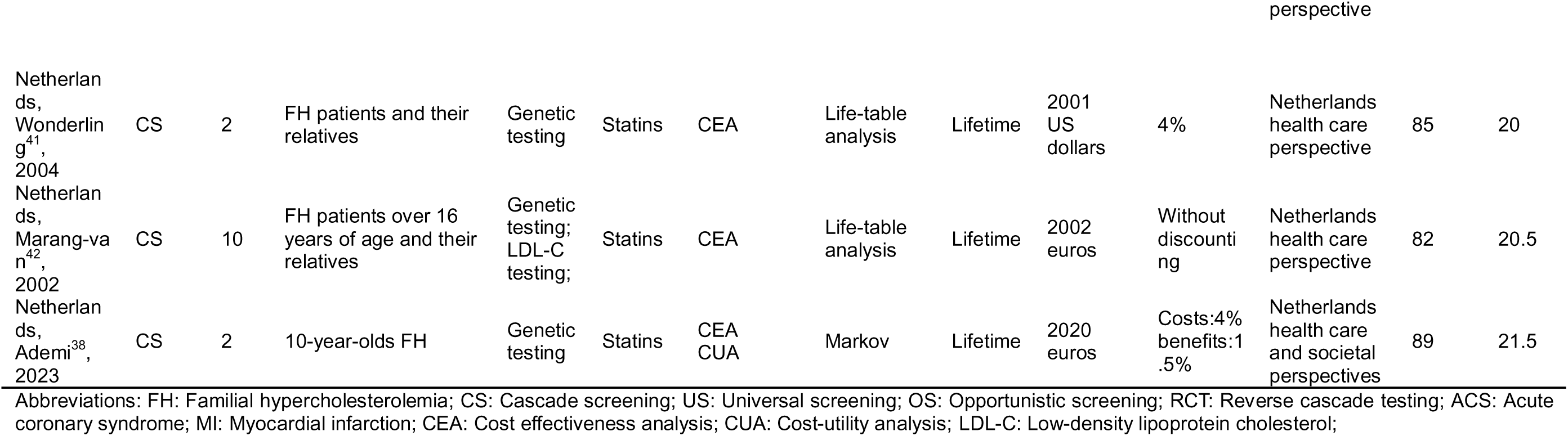
Characteristics and literature quality evaluation of FH screening studies included in the article.

| Country ,<br>Author ,<br>Year , | Strate<br>gy<br>type | Stra<br>tegy<br>num<br>ber | Screening population | Definiti<br>on of<br>FH | Treatm<br>ent | Economic<br>evaluation<br>method | Model | Time<br>horizon | Cost unit | Discount<br>rate | Perspective | QHES<br>score | CHEE<br>RS<br>2022 |
| --- | --- | --- | --- | --- | --- | --- | --- | --- | --- | --- | --- | --- | --- |
| UK,<br>Kerr<sup>30</sup>,<br>2017 | CS | 2 | FH patients and their<br>relatives | Genetic<br>testing | Statins,<br>Ezetimi<br>be | CEA<br>CUA | Markov | Lifetime | 2014-15<br>UK<br>pounds | 3.5% | UK NHS<br>perspective | 87 | 22.5 |
| UK,<br>Crosland<sup>31</sup><br>,<br>2018 | CS | 9 | Patients aged 40-70<br>years with FH<br>and relatives | Genetic<br>testing | Statins | CUA | Decision<br>tree +<br>Markov<br>model | Lifetime | 2015-16<br>UK<br>pounds | 3.5% | UK NHS<br>perspective | 91 | 22 |
| UK,<br>McKay<sup>32</sup>,<br>2018 | US+R<br>CT | 8 | Children 1-2 years old | Genetic<br>testing;<br>LDL-C<br>testing; | Statins | CUA | Decision<br>tree +<br>Markov<br>model | Lifetime | 2017 UK<br>pounds | 3.5% | UK NHS<br>perspective | 86 | 21 |
| UK,<br>Marks<sup>33</sup>,<br>2002 | US;<br>CS;<br>OS; | 5 | General population ;<br>Counseling patients;<br>Inpatients with<br>premature MI; FH<br>patients and their<br>relatives; People aged<br>16-54; | Genetic<br>testing;<br>LDL-C<br>testing; | Statins | CEA | Decision<br>tree +<br>life-table<br>analysis | Lifetime | pounds | costs:6%<br>benefits:1<br>% | UK NHS<br>perspective | 82 | 20 |
| UK,<br>Marks<sup>34</sup>,<br>2003 | US;<br>CS; | 2 | 16-year-olds; Patients<br>aged 16-54 years and<br>their relatives | Genetic<br>testing;<br>LDL-C<br>testing; | Statins | CEA | Decision<br>tree +<br>life-table<br>analysis | 10<br>years | pounds | Not<br>Reported | UK NHS<br>perspective | 80 | 19 |
| UK,<br>Nherera<sup>35</sup>,<br>2011 | CS | 4 | FH patients (50 years<br>old) and relatives (30<br>years old) | LDL-C<br>testing | Statins | CUA | Decision<br>tree +<br>Markov<br>model | Lifetime | 2010-11<br>UK<br>pounds | 3.5% | UK NHS<br>perspective | 89 | 22 |
| Australia,<br>Ademi<sup>45</sup>,<br>2020 | CS | 2 | Ten years olds<br>suspected of having<br>FH | Genetic<br>testing;<br>LDL-C<br>testing; | Statins | CEA<br>CUA | Markov | Lifetime | 2019<br>Australia<br>n dollars | 5% | Australian<br>healthcare<br>system<br>perspective | 91 | 23.5 |
| Australia, Ademi<sup>47</sup>, 2014 | CS | 2 | FH patients and their relatives | Genetic testing; LDL-C testing; | Statins | CEA CUA | Decision tree + Markov | 10 years | 2013 Australian dollars | 5% | Australian health care perspective | 88 | 20.5 |
| Australia, Marquina<sup>36</sup>, 2022 | US | 2 | Population 18–40 years in Australia | Genetic testing | Statins; Ezetimibe | CEA CUA | Decision tree + Markov | Lifetime | 2020 Australian dollars | 5% | Australian healthcare and societal perspective | 93 | 24.5 |
| U.S. Chen<sup>43</sup>, 2015 | CS | 3 | Caucasian male adults | Genetic testing; LDL-C testing; | Statins | CUA | Decision tree + Markov | Lifetime | 2013 US dollars | 3% | U.S. societal perspective | 88 | 21 |
| U.S. Spencer<sup>25</sup>, 2022 | US | 2 | Population-wide (20 and 35 years old) | Genetic testing | Statins | CEA CUA | Decision tree + Markov | Lifetime | 2021 US dollars | 3% | U.S. health care sector perspective | 90 | 22.5 |
| U.S. Jackson<sup>39</sup>, 2021 | CS | 2 | FH+ progenitor population, and the children of the progenitor population, and subsequent off-spring | Genetic testing | Statins, Ezetimibe, PCSK9 | CEA | Simulated family trees; | 30 years | 2018 US dollars | 3% | U.S. health care provider's perspective | 81 | 21.5 |
| Argentina, Araujo<sup>44</sup>, 2023 | US | 2 | Living in Argentina aged 6 years children | LDL-C testing | Statins | CEA CUA | Decision tree | 60 years | US dollars | 5% | Argentine public healthcare system perspective | 84 | 21 |
| Poland, Pelczarska<sup>46</sup>, 2018 | US+CS; OS+CS; | 7 | People who got their first job; 6 years old children; 49/75 years of age after the first ACS/stroke; | Genetic testing; LDL-C testing; | Statins | CEA CUA | Decision tree + Markov | Lifetime | zloty and euros | Costs:5% benefits:3.5% | Poland public payer perspective | 89 | 21 |
| Spanish, Lázaro<sup>37</sup>, 2017 | CS | 2 | FH patients and their relatives | Genetic testing | Statins; Ezetimibe | CEA CUA | Decision tree | 10 years | 2016 euros | 3% | Spanish health system and social perspective | 88 | 21 |
| Spanish, Oliva<sup>40</sup>, 2009 | CS | 2 | Under 60 years old FH and relatives | Genetic testing | Statins | CEA | Life-table analysis | Lifetime | 2005 euros | 3% | Spanish health system | 89 | 21.5 |
| Netherlands, Wonderling<sup>41</sup>, 2004 | CS | 2 | FH patients and their relatives | Genetic testing | Statins | CEA | Life-table analysis | Lifetime | 2001 US dollars | 4% | Netherlands health care perspective | 85 | 20 |
| Netherlands, Marang-van<sup>42</sup>, 2002 | CS | 10 | FH patients over 16 years of age and their relatives | Genetic testing; LDL-C testing; | Statins | CEA | Life-table analysis | Lifetime | 2002 euros | Without discounting | Netherlands health care perspective | 82 | 20.5 |
| Netherlands, Ademi<sup>38</sup>, 2023 | CS | 2 | 10-year-olds FH | Genetic testing | Statins | CEA CUA | Markov | Lifetime | 2020 euros | Costs:4% benefits:1.5% | Netherlands health care and societal perspectives | 89 | 21.5 |

Abbreviations: FH: Familial hypercholesterolemia; CS: Cascade screening; US: Universal screening; OS: Opportunistic screening; RCT: Reverse cascade testing; ACS: Acute coronary syndrome; MI: Myocardial infarction; CEA: Cost effectiveness analysis; CUA: Cost-utility analysis; LDL-C: Low-density lipoprotein cholesterol;

#### Quality Assessment

To ensure the inclusion of literature adhering to high-quality research methods and reporting standards, a comprehensive quality assessment of the selected studies was conducted. Two distinct tools were employed for this purpose: the Quality of Health Economic Studies (QHES) instrument and the Consolidated Health Economic Evaluation Reporting Standards (CHEERS) 2022.

#### QHES instrument Evaluation

The Quality of Health Economic Studies (QHES) scale, a well-established instrument consisting of 16 key criteria, served as the primary tool for assessing the methodological rigor and overall quality of the selected studies.^21^ Each criterion was diligently evaluated, with scores ranging from 0 to 100, offering a comprehensive overview of the studies’ methodological soundness. Subsequently, the total scores were categorized into levels indicative of study quality: high quality (75-100), moderate quality (50-74), low quality (25-49), and very low quality (0-24).^22^ See Appendix 4 for details of the QHES score.

#### CHEERS 2022 Evaluation

CHEERS 2022 served as a quantitative scoring system for the systematic review, encompassing 28 scoring criteria across seven critical domains, including title, abstract, introduction, methods, results, discussion, and others.^23^ Each criterion was assigned specific scoring rules: complete reporting earned 1 point, partial reporting earned 0.5 points, and non-reporting received 0 points. The maximum total score was 28 points, with detailed scoring specifics available in the Appendix 3.^24^

### 2. Meta-Analysis

#### Eligibility for meta-analysis

Following the systematic review, we conducted a meta-analysis to synthesize the economic evidence. Apparently, total articles were eligible for evidence synthesis since they were dealing with heterogeneous contexts. We applied some eligibility criteria to select articles for evidence synthesis that generated sensible outcomes. First, the included studies were consistent in terms of the screening strategies, such as cascade or universal screening. Second, we ensured the outcomes of included studies were consistent, such as CE, Life Years Gained, or Quality-Adjusted Life Years. Finally, the included studies applied the same perspective, whether it be a healthcare system or a societal perspective.

#### Evidence synthesis Cost

We extracted cost values from eligible studies for the meta-analysis. Due to the heterogeneity in the use of currencies and years across different articles, we converted all costs to 2023 US dollars for calculation to ensure consistency. For those without reported confidence intervals, we calculated the intervals using a range of ±50%. The types of costs are shown in Appendix 2.

#### Effectiveness

Meta-analysis of health outcomes resulting from FH screening involved categorization into four distinct classes: deaths averted, adverse events averted, Life Years Gained and Quality-Adjusted Life Years. Extracted by different result categories in Appendix 5. Results lacking reported confidence intervals were handled conservatively, with a ±50% adjustment to address potential variability in the numerical values. Additionally, Spencer et al. reported results for 20 and 35-year-old cohorts, combining averages of effectiveness for both age groups. ^25^

#### Incremental benefit calculation

Integration of health outcomes and incremental costs was performed to determine the ICER. The total incremental costs and total effectiveness were computed, and the ICER was derived by dividing the synthesized cost by synthesized health outcomes (i.e., QALYs, LYGs). Confidence intervals for ICERs were obtained using the Delta method. The Delta method involves calculating the variance of the ratio. Utilizing this variance of the ratio, the standard error is computed, subsequently allowing the derivation of a 95% confidence interval for ICER values. This method ensures a robust statistical approach to ascertain the uncertainty surrounding the cost-effectiveness estimates (Table 3).

#### Total Net Health Benefit

To facilitate a comprehensive evaluation of different FH screening groups, we employed the Comparative efficiency research (COMER).^26^ Initially, we utilized this method to calculate the Net Health Benefit included in the study. NHB is a comprehensive indicator in health economics evaluations that takes into account both costs and benefits. The positive or negative sign and magnitude of its results provide an intuitive reflection of the efficiency levels of health projects. Subsequently, by taking the reciprocal of the variance of each study’s NHB and applying weighting, we calculated the Total Net Health Benefit (TNHB) for different FH screening groups. The detailed calculation methodology for COMER can be found in the Appendix 6. Furthermore, to enhance the granularity of our analysis, we calculated the incremental costs and incremental effects of each group of weights according to the weight value (Table3).

### 3. Aggregate DCEA

In our study, we conducted an aggregate Distributional Cost-Effectiveness Analysis based on the original cost-effectiveness analysis results of FH screening. This was undertaken to comprehensively assess how the health impacts and costs post FH screening are distributed among population subgroups and to estimate the fairness impact of FH screening strategies. The aggregate DCEA analysis provides detailed information on the degree and direction of the fairness impact of FH screening, offering policymakers a robust reference. The following are the steps we took:

#### Health Distribution

In the absence of interventions, the baseline health level of the population was established. The baseline distribution of population health and the distribution of health opportunity costs were derived from the work of Love-Koh et al.^27,28^ Their study delineated the distribution of quality-adjusted life expectancy (QALE) at birth and the corresponding distribution of medical opportunity costs. To measure socio-economic poverty, the study employed the Index of Multiple Deprivation (IMD), categorizing the population into five groups, IMD1 to IMD5, where IMD1 represents the most impoverished areas, and IMD5 represents the most affluent areas.^19^ The Baseline distribution of health for IMD1 and IMD5 was 63.21 and 75.00, respectively, while the health opportunity costs distribution was 26% and 14%, respectively (Appendix 7). Subsequently, we incorporate the Net Health Benefit resulting from FH screening into the baseline population health distribution, yielding the health distribution after FH screening.

#### Opportunity cost relevant Net Health Benefit

For each Index of Multiple Deprivation (IMD) group, Incremental NHB is calculated, and these values are subsequently consolidated. The NHB is determined by subtracting opportunity costs from the overall health gains. The threshold for health opportunity costs is set at the upper limit of the willingness-to-pay threshold, specifically the maximum willingness-to-pay threshold utilized in the study.

The calculation formula is as follows: Aggregated incremental net health benefit per quintile:

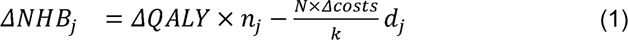

where *ΔNHBj* represents the incremental net health benefit for the *j^th^* group; *ΔQALY* denotes the incremental health gains from FH screening; *nj* is the number of individuals screened in the *j^th^* group, *N* is the total number of screenings, *K* is the threshold for health opportunity costs, *dj* is the opportunity cost ratio for the *j^th^* group, and *Δcosts* refer to the incremental costs of screening for individuals.

#### Equally distributed equivalent health

Equally distributed equivalent health (EDEH) is the equitable weighted average of health distribution. By calculating the post-decision EDEH and subtracting it from the baseline EDEH, the incremental EDEH is obtained. We computed the EDEH using the Atkinson inequality index and the average health level in the distribution. The Atkinson index represents inequality in the distribution, and the Atkinson Inequality Aversion Parameter (IAP) estimated according to Robson was 10.95.^29^ Using the formulas for Atkinson index of inequality and EDEH based on the Atkinson index, where N is scaled based on the number of screened patients:

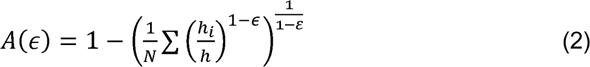

where *A(ε)* is the Atkinson Inequality Index, where *N* represents the total number of screenings, *hi* is the health level of each group, *h* is the average health level of the population, and *E* is a constant aversion to relative inequality levels.

#### *EDEH* based on Atkinson index

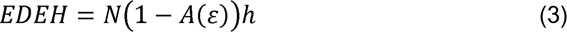

where *EDEH* refers to the average level of per capita health, *A(ε)* is the Atkinson Index, *h* is the average health level of the population, and *N* is the total number of screenings.

#### EDEH-NHB

We computed the incremental EDEH and incremental NHB to assess changes in health distribution. The impact on health equity is evaluated by calculating the difference between EDEH and NHB. A positive value indicates a reduction in health inequality, while a negative value indicates an increase in inequality.^19^

## RESULTS

### 1. Literature Search Results

#### Inclusion of Literature

Through systematic searches of databases such as PubMed (47 articles), Web of Science (109 articles), and Embase (33 articles), we identified a total of 189 articles. After importing these articles into EndNote 20, 38 duplicates were removed, and 98 articles were excluded based on exclusion criteria, resulting in the final inclusion of 18 articles. Subsequently, an additional article was included, bringing the total to 19 articles for the final analysis. Please refer to Figure 1 for a detailed flowchart of the literature selection process (Figure 1).

**Figure 1.**
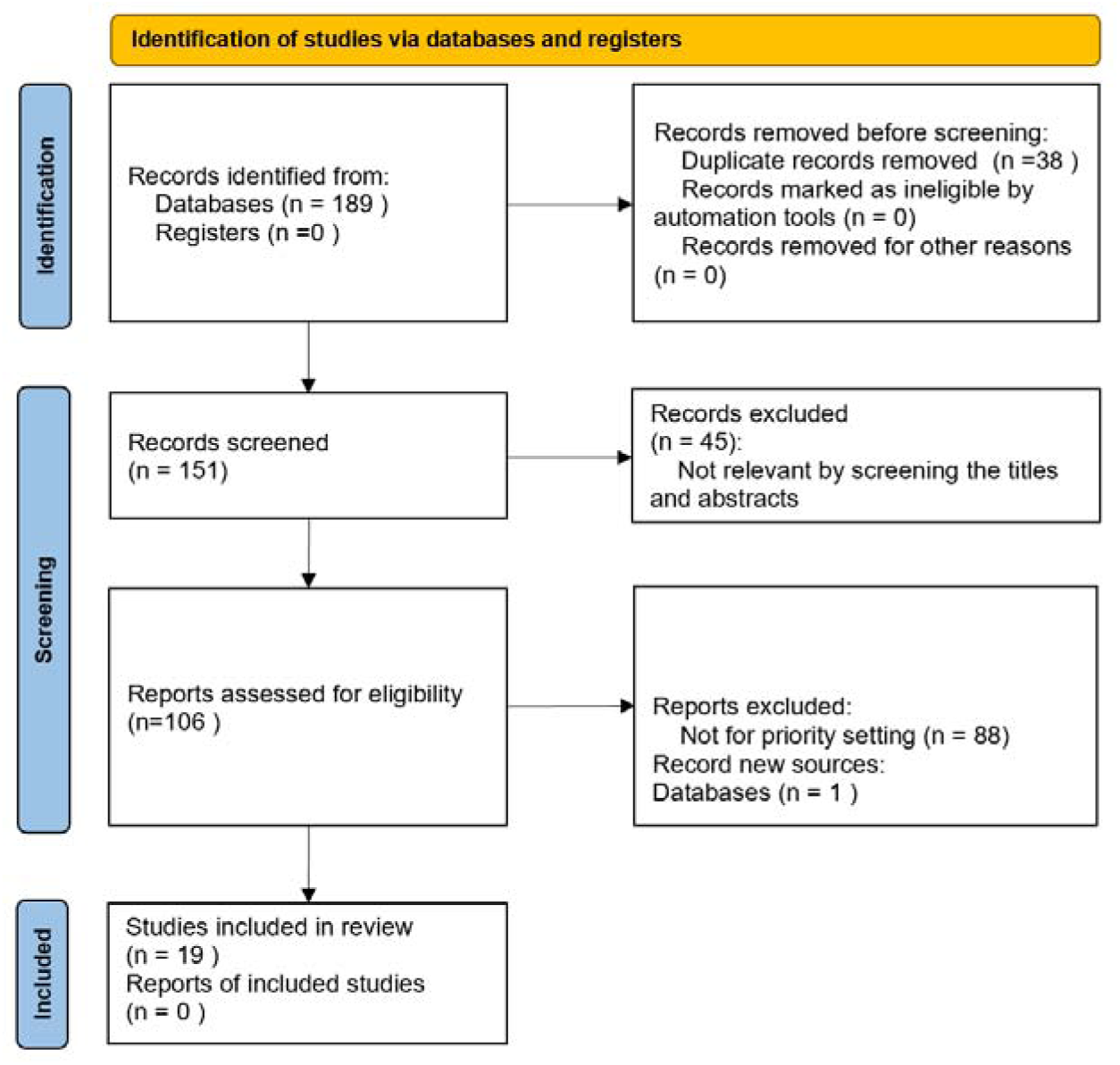
Preferred Reporting Items for Systematic Reviews and Meta-Analyses Flow Diagram Illustrating the Study Selection Process

#### Quality Assessment outcomes

The average QHES evaluation score stands at 87, with a score range spanning from 80 to 93. This signifies that the entirety of the selected literature attains a high-quality standard, achieving a 100% level of excellence. Furthermore, aligning with the CHEERS 2022 assessment, these articles garnered scores within the range of 19 to 24.5, with an average score of 21 for the 19 articles, once again affirming the exceptional quality of the chosen literature (Table 1). Throughout the scrutiny of each economic evaluation study, the employed methods and models demonstrated clear and transparent characteristics. They were elucidated with detailed explanations of the selected model’s temporal framework. Despite variances in FH screening strategies across diverse countries, all articles employed explicit language to articulate comparator strategies. Moreover, they provided explicit and comprehensive reports of measurement results, establishing a robust foundation for the comparability of research outcomes.

#### Characteristics of included studies

This study involves a comprehensive analysis of 19 articles, covering different geographical focuses, with 32% conducted in the United Kingdom,^30–35^ and 63% are concentrated in Europe. Regarding the analytical perspective, 79% of the studies focused solely on healthcare, considering direct medical costs such as screening and treatment expenses,16% concurrently considered healthcare and societal perspectives.^36–38^ A substantial 89% discounted costs and health outcomes. Among the discounted studies, 82% employed identical discount rates for costs and benefits, with cost discount rates ranging from 3% to 6% and benefit discount rates ranging from 1% to 5%.

In the literature, 32% employed cost-effectiveness analysis,^33,34,39–42^ while 21% used cost-utility analysis with QALY as the health outcome.^31,32,35^ The modeling approaches varied, encompassing Markov models, decision trees, life-table analysis, simulated family trees, and combinations thereof. Markov models were the most prevalent, constituting over half of the studies, often reporting health state numbers ranging from 3 to 14. Notably, a unique U.S. study incorporated simulated family trees as part of its modeling approach.^39^ In Appendix 1 and Table 1, we have presented a detailed display of the review results.

Among the FH screening studies, 90% reported ICER values below their respective country’s willingness-to-pay thresholds, indicating cost-effectiveness. However, two U.S. studies introduced divergent findings.^43^ Specifically, genomic screening was deemed not cost-effective at the current U.S. willingness-to-pay threshold.^25^

To fortify the robustness of the models, 95% of the studies conducted sensitivity analyses. Among them, 53% utilized probabilistic sensitivity analysis, simulating cost-effectiveness probabilities and drawing cost-effectiveness acceptability curves (CEACs) based on thresholds. It is worth noting that, with variations in the willingness-to-pay thresholds, there are significant differences in the probability of cost-effectiveness for the screening. For instance, in the U.S. population-wide genomic screening, for individuals aged 20, the cost-effective probabilities of FH screening were 1%, 38%, and 81% at QALY thresholds of $50000, $100000, and $150000, respectively. For those aged 35, the corresponding probabilities were 0%, 14%, and 57%.^25^ (Table 2)

**Table 2.**
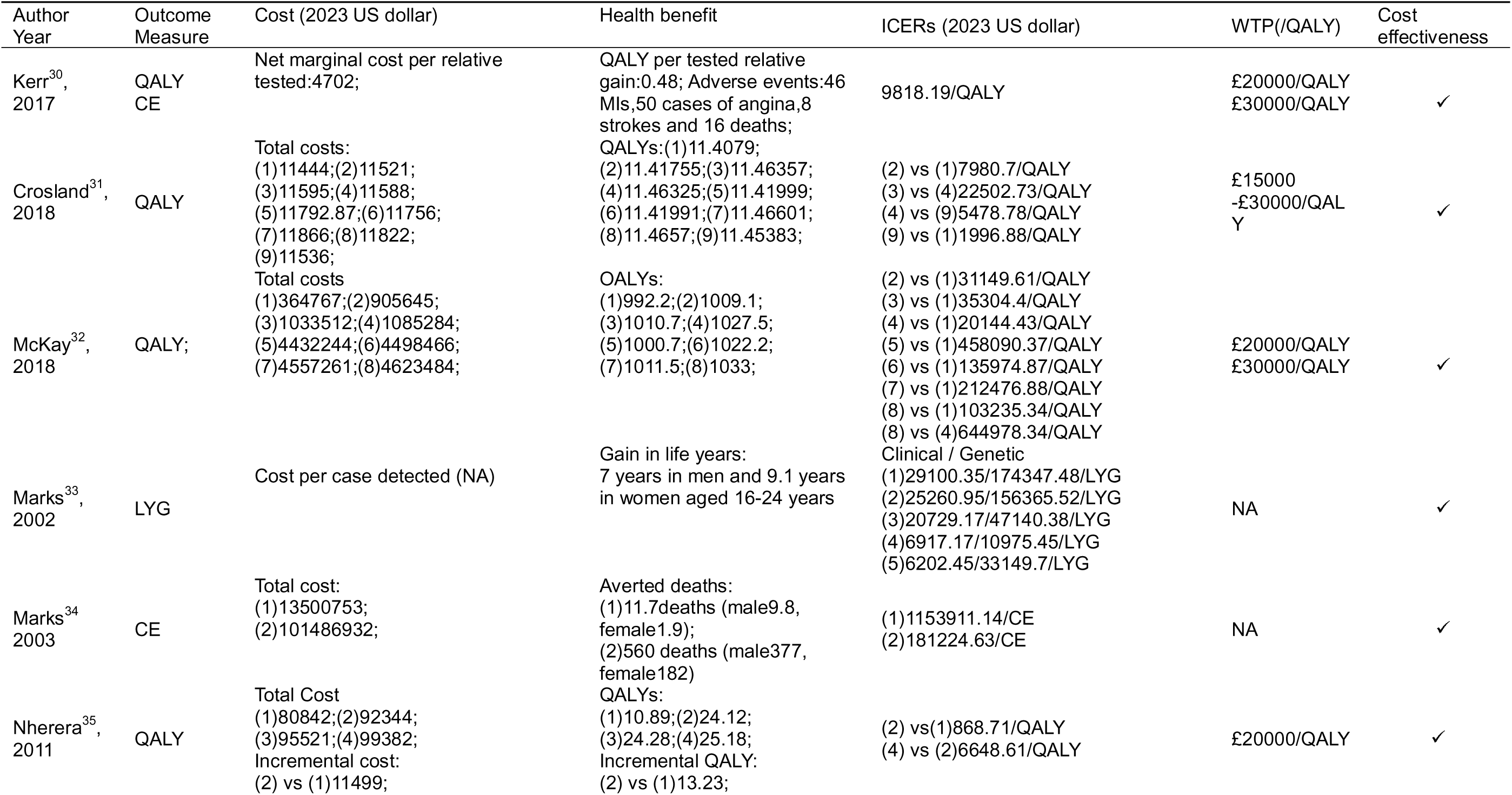
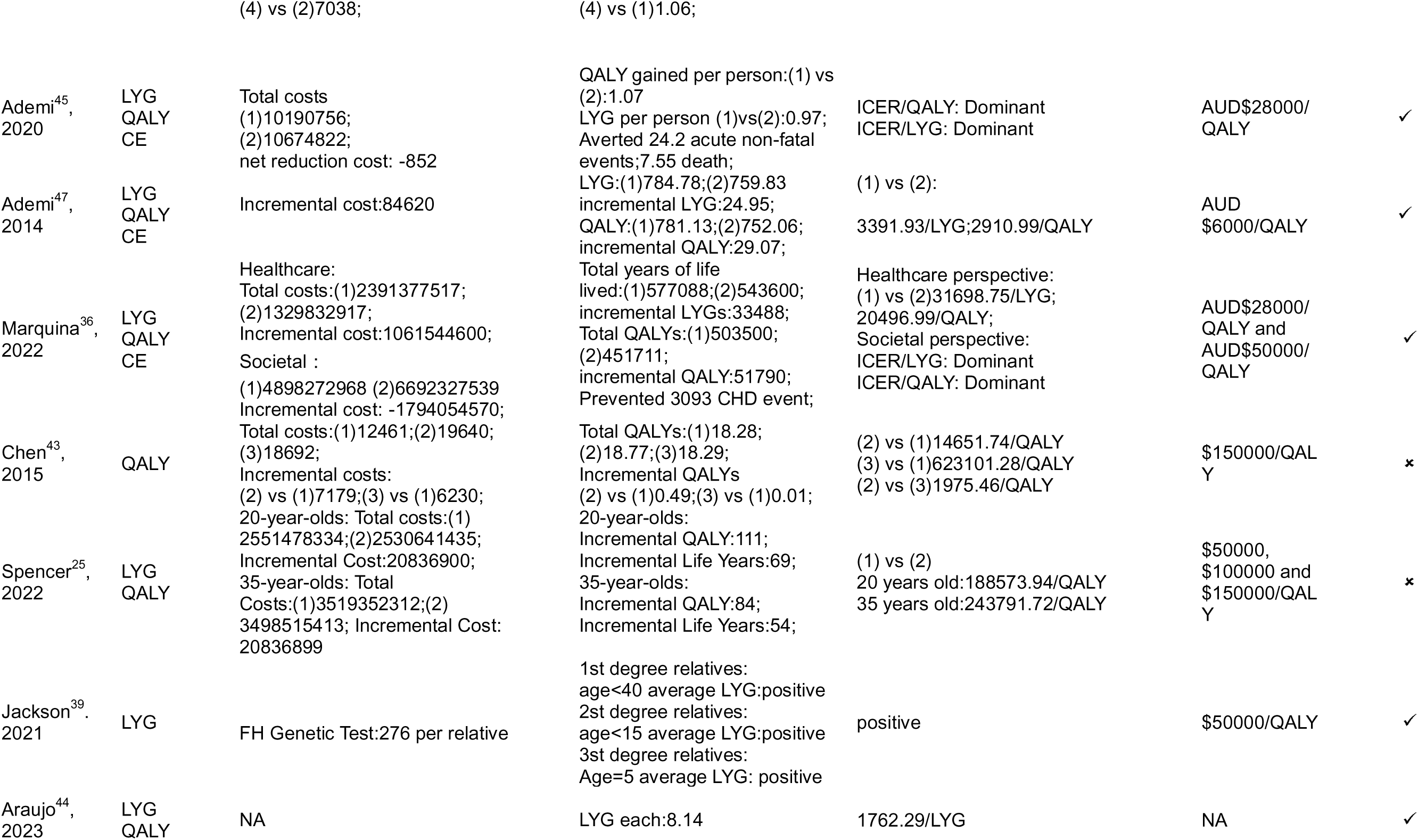
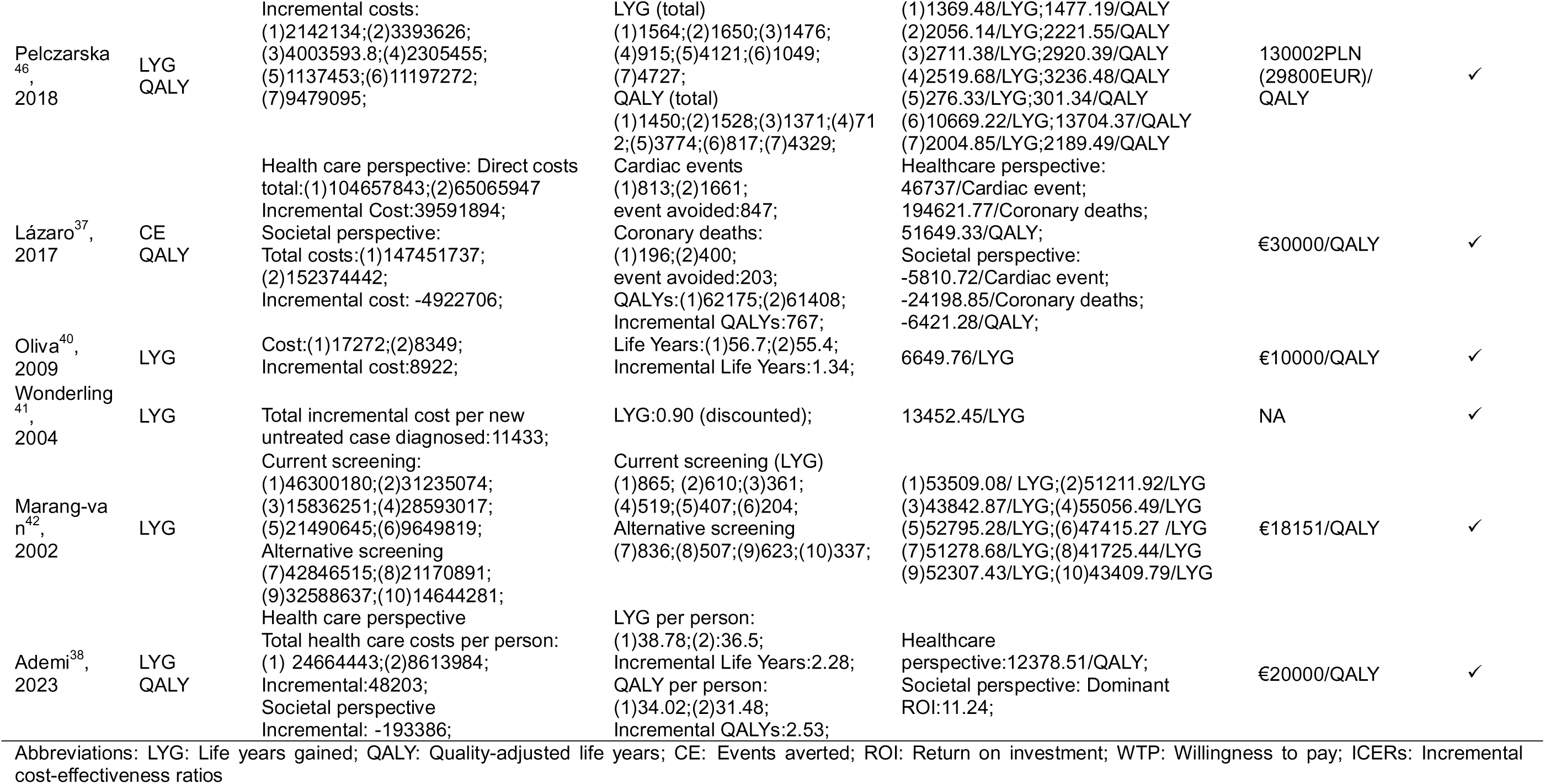
Summary of economic results is included in the article.

#### FH screening strategies

##### Child Screening

The atherosclerosis process initiates in childhood, and child screening has demonstrated favorable cost-effectiveness in select countries such as the UK, Australia, Argentina, and the Netherlands. McKay et al. conducted universal screening of 1-2-year-old children in the UK, followed by reverse cascade testing. The findings indicated that this strategy was economically viable within the UK’s willingness-to-pay threshold.^32^ In Argentina, a probabilistic model assessed the expected cost-effectiveness of universal FH screening for 6-year-old children, revealing it as a highly cost-effective health technology.^44^ Similar studies were conducted in Australia and the Netherlands. Ademi et al^38,45^ the economic aspects of cascade screening for 10-year-old children from the perspectives of the Australian public healthcare system and Dutch healthcare and society, respectively. Results consistently showed that cascade screening for 10-year-old children was cost-effective compared to standard care in both Australia and the Netherlands.

##### Several Age-Groups

Economic evaluations of FH screening for various age groups were conducted globally. In the United States, a study on comprehensive genomic FH screening found improved cost-effectiveness for screening younger patient cohorts compared to older ones.^25^ An Australian study conducted a cost-effectiveness assessment of genomic screening for young individuals with FH. Subgroup analysis revealed that narrowing the screening age range from 18-40 years to 18-25 years resulted in an increased cost per Quality-Adjusted Life Year.^36^ Another Australian study focusing on cost-effectiveness analysis of cascade screening for FH in children conducted subgroup analyses for different age groups. The results revealed that, compared to 18-year-olds, screening 10-year-old children for FH and initiating statin therapy immediately remained cost-saving.^45^

##### Cascade screening and optimization

The cost-effectiveness of cascade screening has been substantiated in numerous countries, demonstrating commendable efficiency. Some nations have integrated cascade screening with different case identification methods to determine the most cost-effective screening strategy. These methods include searching electronic health records, utilizing various clinical assessment standards,^31^ screening identified cases separately based on genetic testing and cholesterol testing,^35^ combining genetic testing and cholesterol testing but distinguishing the order,^32^ The results underscore that incorporating these diverse case identification methods surpasses the cost-effectiveness of standalone cascade testing.^31^

##### Strategies combination

Recognizing the complementarity of these strategies, some countries are exploring combinations for a more comprehensive FH screening approach. In Poland, researchers combined Universal Screening followed by Cascade Screening for different populations or Opportunistic Screening followed by Cascade Screening for clinically or genetically diagnosed high-risk populations. Results from the evaluation of seven strategies indicated that screening patients with acute coronary syndrome (ACS) under 55/65 years of age using clinical criteria emerged as the most cost-effective strategy. From the perspective of public payers, the most acceptable solution for introducing FH screening might be a combination of multiple strategies.^46^

### 2. Meta-Analysis Results

The cost-effectiveness analysis of the literature revealed that 90% of FH screening studies were deemed cost-effective (Table 2).The meta-analysis excluded literature that did not provide specific cost or outcome values^33,39,44^ and literature where strategies were not comparable.^31,32,35,42,43,46^ The final synthesis of results included eight studies on cascade screening^30,34,37,38,40,41,45,47^ and three studies on Universal Screening.^25,34,36^ These studies considered outcome measures such as QALY, LYG, adverse events averted, and deaths averted, resulting in the eventual synthesis of seven distinct groups (Table 3).

**Table 3.** Synthesis of cost-effectiveness analysis results and COMER outcomes.

|  | Study | Incremental costs<br>(2023 US dollar) | Incremental<br>effects | Total ICER<br>(2023 US dollar) | NHB<br>(2023 US dollar) | NHB<0(%) | Weight (%) |
| --- | --- | --- | --- | --- | --- | --- | --- |
| CS(QALY) | Kerr<sup>30</sup>,2017 | 4702.79 | 0.48 | 49630(37223 to 62038) | 19648 | NHB>0 | 0.4734 |
|  | Ademi<sup>45</sup>,2020 | -852.31 | 1.07 |  | 23369 | NHB>0 | 0.3346 |
|  | Ademi<sup>47</sup>,2014 | 84620.28 | 29.07 |  | 57823 | NHB>0 | 0.0547 |
|  | Lázaro<sup>37</sup>,2017 | 39591894.47 | 767 |  | 547516 | NHB>0 | 0.0006 |
|  | Ademi<sup>38</sup>,2023 | 31369.18 | 2.53 |  | 36563 | NHB>0 | 0.1367 |
| | | $\Sigma\omega * c=34991$ | $\Sigma\omega * e=2.99$ | | TNHB=25614(19210 to 32017) | TNHB>0 | $\Sigma w=1$ |
| CS(LYG) | Ademi<sup>45</sup>,2020 | -852.31 | 0.97 | 4451(3338 to 5564) | 21264 | NHB>0 | 0.225 |
|  | Ademi<sup>47</sup>,2014 | 84620.28 | 24.95 |  | 37634 | NHB>0 | 0.0718 |
|  | Oliva<sup>40</sup>,2009 | 8922.69 | 1.34 |  | 17108 | NHB>0 | 0.3477 |
|  | Wonderling<sup>41</sup>,2004 | 11433.06 | 0.9 |  | 20539 | NHB>0 | 0.2412 |
|  | Ademi<sup>38</sup>,2023 | 31369.18 | 2.28 |  | 29851 | NHB>0 | 0.1142 |
| | | $\Sigma\omega * c=15331$ | $\Sigma\omega * e=2.95$ | | TNHB=21801(16351 to 27251) | TNHB>0 | $\Sigma w=1$ |
| CS<br>(adverse<br>events<br>averted) | Kerr<sup>30</sup>,2017 | 4702.79 | 104 | 40603(30452 to 50753) | 5271321 | NHB>0 | 0.0092 |
|  | Ademi<sup>45</sup>,2020 | -852.31 | 24.2 |  | 510117 | NHB>0 | 0.9795 |
|  | Lázaro<sup>37</sup>,2017 | 39591894.47 | 847 |  | 4734156 | NHB>0 | 0.0114 |
| | | $\Sigma\omega * c=449450$ | $\Sigma\omega * e=34.29$ | | TNHB=601825(451368 to 752281) | TNHB>0 | $\Sigma w=1$ |
| CS (deaths<br>averted) | Kerr<sup>30</sup>,2017 | 4702.79 | 16 | 179369(134526 to 224211) | 806993 | NHB>0 | 0.0377 |
|  | Marks<sup>34</sup>,2003 | 101486932.5 | 560 |  | -64766053 | NHB<0 | 0.000005 |
|  | Ademi<sup>45</sup>,2020 | -852.31 | 7.55 |  | 159735 | NHB>0 | 0.9633 |
|  | Lázaro<sup>37</sup>,2017 | 39591894.47 | 203 |  | -28968295 | NHB<0 | 0.00003 |
| | | $\Sigma\omega * c=1110$ | $\Sigma\omega * e=7.88$ | | TNHB=182905(137178 to 228631) | TNHB>0 | $\Sigma w=1$ |
| US(QALY) | Marquina<sup>36</sup>,2022 | 1061544599.77 | 51790 | 20860(15645 to 26075) | 854219290 | NHB>0 | 0.00004 |
|  | Spencer<sup>25</sup>,2022 | 20836899.42 | 97.5 |  | -5599989 | NHB<0 | 0.99996 |
| | | $\Sigma\omega*c=20881624$ | $\Sigma\omega * e=99.72$ | | TNHB=-5563039(-6953798 to -4172279) | TNHB<0 | $\Sigma w=1$ |
| US (deaths averted) | Marks<sup>34</sup>,2003 | 13500753.09 | 11.7 | 832917(624687 to 1041146) | -12733549 | NHB<0 | 0.99998 |
|  | Marquina<sup>36</sup>,2022 | 1061544599.77 | 1279 |  | -1014233111 | NHB<0 | 0.00002 |
| | | $\Sigma\omega * c=13665924$ | $\Sigma\omega * e=11.9$ | | TNHB=-12891385(-16114231 to -9668539) | TNHB<0 | $\Sigma w=1$ |
| US(LYG) | Marquina<sup>36</sup>,2022 | 1061544599.77 | 33488 | 32262(24197 to 40328) | 177210008 | NHB>0 | 0.004 |
|  | Spencer<sup>25</sup>,2022 | 20836899.42 | 61.5 |  | -11225925 | NHB<0 | 0.996 |
| | | $\Sigma\omega * c =24996554$ | $\Sigma\omega * e =195.1$ | | TNHB=-10472757(-13090946 to -7854568) | TNHB<0 | $\Sigma w=1$ |

Abbreviations: CS: Cascade screening; US: Universal screening; $\omega$ : weigh; $\Sigma\omega*c$ : The weighted sum of costs; NHB: Net health benefit; TNHB: Total health benefit; LYG: Life years gained; QALY: Quality-adjusted life years; CE: Events averted.

#### Cost-Effectiveness of Cascade Screening

For studies using QALY as the outcome measure,^30,37,38,45,47^ the synthesized results showed a sum Incremental Cost of $39711734, a total QALYs gain of 800, and a calculated ICER per QALY of $49630. For studies using LYG as the outcome measure,^38,40,41,45,47^ the synthesized results showed a sum Incremental Cost of $135493, sum LYG per person of 30.44, and a calculated ICER per LYG of $4451.Studies using adverse events averted as the outcome measure indicated a sum Incremental Cost of $39595745, sum adverse events averted was 975.2, and the calculated ICER was $40603. For studies using deaths averted as the outcome measure,^30,34,37,45^ the sum Incremental Cost was $141082678, sum deaths averted was 786.55, and the calculated ICER was $179369.

#### Cost-Effectiveness of Universal Screening

Similarly, for studies using LYG and QALY as the outcome measures,^25,36^ The synthesized results showed a Sum Incremental Cost of $1082381499, Sum LYG per person of 33549.5, and a calculated ICER per LYG of $32262. The Sum QALY per gain was 51877.5, and the calculated ICER per QALY was $20860. For studies using deaths averted as the outcome measure,^34,36^ the synthesized results showed a Sum Incremental Cost of $1075045353, Sum deaths averted was 1290.7, and the calculated ICER was $832917.

#### TNHB

Analyzing the COMER results, we present the synthesized data for Net Health Benefit across seven FH screening groups with distinct characteristics. The TNHB for the four cascade screening groups consistently exhibited positive values, whereas for the three universal screening groups, TNHB consistently showed negative values. Specifically, for cascade screening, the TNHB for QALYs was $25614, for LYG was $21801, for adverse events averted was $601825 and for deaths averted was $182905. In contrast, for universal screening, the TNHB for QALYs was -$5563039, for LYG was -$10472757 and for deaths averted was -$12891385. The results are summarized in Table 3.

### 3. Aggregate DCEA

#### Average Baseline EDEH

In this study, the mean Quality-Adjusted Life Years per individual stood at 69.72, corresponding to an Equivalent Disposable Income per Equivalent Adult (EDEH) of 68.32, given an Atkinson Individual Average Propensity (IAP) of 10.95.

#### DCEA Calculation

Given the heterogeneity in costs, health outcomes, and opportunity cost thresholds across the 19 articles, we opted to conduct aggregate Distributional Cost-Effectiveness Analysis calculations on articles reporting complete data, resulting in 6 final publications (Table 4).

**Table 4.**
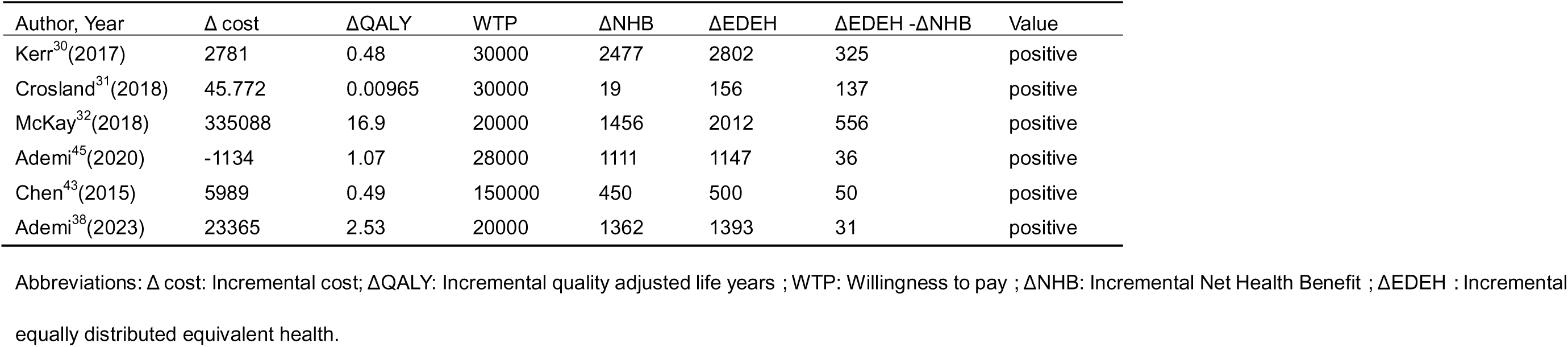
The impact of FH screening strategies on health equity.

| Author, Year | $\Delta$ cost | ΔQALY | WTP | ΔNHB | ΔEDEH | ΔEDEH -ΔNHB | Value |
| --- | --- | --- | --- | --- | --- | --- | --- |
| Kerr<sup>30</sup>(2017) | 2781 | 0.48 | 30000 | 2477 | 2802 | 325 | positive |
| Crosland<sup>31</sup>(2018) | 45.772 | 0.00965 | 30000 | 19 | 156 | 137 | positive |
| McKay<sup>32</sup>(2018) | 335088 | 16.9 | 20000 | 1456 | 2012 | 556 | positive |
| Ademi<sup>45</sup>(2020) | -1134 | 1.07 | 28000 | 1111 | 1147 | 36 | positive |
| Chen<sup>43</sup>(2015) | 5989 | 0.49 | 150000 | 450 | 500 | 50 | positive |
| Ademi<sup>38</sup>(2023) | 23365 | 2.53 | 20000 | 1362 | 1393 | 31 | positive |

Abbreviations: $\Delta$ cost: Incremental cost; ΔQALY: Incremental quality adjusted life years ; WTP: Willingness to pay ; $\Delta$ NHB: Incremental Net Health Benefit ; $\Delta$ EDEH : Incremental equally distributed equivalent health.

In the UK’s 2017 cascade screening, with an opportunity cost threshold of £30000/QALY, the incremental NHB for FH cascade screening compared to no screening was 2477 QALYs. The post-decision EDEH was 2802 QALYs, with a difference of 325 QALYs, indicating a reduction in inequality.^30^

In the UK’s 2018 cascade screening, with an opportunity cost threshold of £30,000/QALY, the incremental NHB for cascade screening of FH patients compared to no screening was 19 QALYs. The post-decision EDEH was 156 QALYs, with a difference of 137 QALYs, indicating a reduction in inequality.^31^

In the UK’s universal screening plus reverse cascade screening, with an opportunity cost threshold of £20000/QALY, the incremental NHB for cholesterol-based cascade screening of FH patients compared to no screening was 1456 QALYs. The post-decision EDEH was 2012 QALYs, with a difference of 556 QALYs, indicating a reduction in inequality.^32^

In Australia’s cascade screening, with an opportunity cost threshold of AU$28000/QALY, the incremental NHB for FH cascade screening compared to no screening was 1111 QALYs. The post-decision EDEH was 1147 QALYs, with a difference of 36 QALYs, indicating a reduction in inequality.^45^

In the US’s cascade screening, with an opportunity cost threshold of $150000/QALY, the incremental NHB for adding medication adherence to cholesterol screening of FH patients compared to cholesterol screening alone was 450 QALYs. The post-decision EDEH was 500 QALYs, with a difference of 50 QALYs, indicating a reduction in inequality.^43^

In the Netherlands’ cascade screening, with an opportunity cost threshold of €20000/QALY, the incremental NHB for cascade screening of children with FH compared to no screening was 1362 QALYs. The post-decision EDEH was 1393 QALYs, with a difference of 31 QALYs, indicating a reduction in inequality.^38^

#### CEA and DCEA Integration

Through cost-effectiveness analysis, FH screening was found to be cost-effective in majority of cases, with numerous studies identifying the most cost-effective options. Building upon these CEA analyses, DCEA calculations were performed, examining the impact on the distribution of health after allocating resources to FH screening. Across the six analyzed publications, all EDEH-NHB values were positive, indicating that the FH screening strategy, as an intervention measure, is capable of reducing inequality, improvement in population health and health equity, and an increase in social welfare. Please refer to Table 4 for detailed DCEA results.

## DISCUSSION

This study represents the first-ever global comprehensive economic evidence synthesis on FH screening, addressing a crucial void in the existing literature. The absence of systematic evaluations in numerous countries prompted our investigation, which conducted an exhaustive literature search and synthesized evidence, contributing nuanced economic evaluation outcomes and health impact analyses associated with FH screening. The amalgamation of international experiences and evaluation outcomes paints an optimistic picture, suggesting the economic feasibility of FH screening. This effort not only offers technical guidance for FH screening economic evaluations on a global scale but also establishes a robust foundation for future economic evaluations in this domain.

Upon closer scrutiny of the economic evaluations of FH screening, a significant heterogeneity emerges, necessitating a comprehensive consideration of multiple factors. Firstly, the analytical perspective stands out as a key consideration. Most studies predominantly adopted a payer perspective, neglecting the comprehensive account of productivity losses. Even more, we need to calculate the return on investment of FH screening from a broader perspective, as the Netherlands has done.^38^ Secondly, the choice of models emerges as a critical consideration. While Markov models are widely applied, the utilization of composite models, combining decision tree models with other approaches, can more comprehensively address the actual complexities associated with FH screening. This enhancement in model reliability ensures a more accurate representation of the economic implications. Thirdly, flexibility in selecting time models based on practical exploration is paramount. The majority of studies focus on a lifetime horizon, but considering different tracking periods, such as 10 years, 30 years, and 60 years, enhances the adaptability of models to varying research needs, thereby increasing their practicality.

The demonstrated cost-effectiveness of cascade screening in an increasing number of countries highlights its importance. However, the exploration of cascading through multiple generations remains an important avenue for investigation. A study in the United States, simulating approximately 6 million individuals using the Simulation of Family Tree, revealed that beyond first and second-degree relatives, cascade screening is not cost-effective.^39^ While many countries have conducted cascade screening and demonstrated its economic benefits, only a study in the United Kingdom has applied reverse cascade screening, proving its economic effectiveness after universal screening for children. This underscores the importance of future discussions on the strategic integration of reverse cascade screening for FH in children.^32^

The crucial importance of determining the cost-effectiveness of health technology, particularly in the context of publicly funded healthcare insurance systems, cannot be overstated. In many countries, the cost-effectiveness of FH screening remains uncertain, emphasizing the need to establish this before considering large-scale implementation.^48^ Our study’s promising outlook, revealing that FH screening is cost-effective in 90% of cases, underscores its potential contribution to the overall efficiency of healthcare systems. Policymakers can leverage these findings to enhance the financial sustainability of healthcare systems, ultimately contributing to improved patient health.

Analyzing the impact of FH screening on population equity, we found that FH screening can simultaneously improve health equity and elevate health levels in various circumstances. In many countries, where health disparities persist, our results provide a positive perspective on the potential of FH screening, particularly for resource-limited and impoverished regions. The FH screening intervention is poised to bring about significant health outcomes in these countries, thereby contributing to the improvement of societal fairness and equality.

Turning our attention to treatment options for FH, a diverse array of options, particularly in reducing low-density lipoprotein cholesterol (LDL-C) levels, is available. Statins, widely used in FH treatment, effectively reduce LDL-C levels by inhibiting cholesterol synthesis enzymes. However, for FH patients requiring high-dose statin treatment yet intolerant to its side effects, PCSK9 inhibitors may emerge as a crucial alternative.^49^ Although PCSK9 inhibitors demonstrate remarkable effectiveness in lowering LDL-C levels, their cost-effectiveness in patients with heterozygous familial hypercholesterolemia does not meet the generally accepted incremental cost-effectiveness threshold.^50^ The potential cost-effectiveness of screening plus PCSK9 treatment approaches remains unclear. It is imperative to consider them in the broader context of screening and treatment strategies in future economic evaluations.

Precision public health, aiming to provide the right intervention to the right population at the right time, is a continually evolving field. The cost-effectiveness of genetic testing and cholesterol testing in FH screening economic evaluations varies between countries, influencing economic outcomes.^48^ In a study conducted in the UK, all DNA-based methods were shown to be cost-effective compared to cholesterol-only methods.^35^ However, in some US-based studies, the cost-effectiveness of genetic testing is challenged by the high costs associated with it.^25,43^ This underscores the necessity of careful consideration of the complexities and factors involved in precision interventions. When genetic testing costs are prohibitively high, plasma cholesterol level screening may be the more appropriate intervention. Additionally, lowering the cost of genomic testing or integrating FH screening into broader multiphasic screening programs may offer more efficient intervention strategies, promoting the implementation of precision public health strategies.

## CONCLUSION

Our research offers valuable insights into the economic evaluation of screening strategies for FH, revealing substantial differences among various countries in the economic assessment process. Despite variations in the economic evaluation across different studies, the attainment of a 90% FH screening rate demonstrates cost-effectiveness. Importantly, the implementation of these screening strategies extends beyond mere cost-effectiveness. Our findings highlight that they not only enhance population health but also contribute to the reduction of inequality, promoting health equity. This dual impact presents a positive outlook for the economic evaluation of FH screening in many countries.

## Data Availability

All data produced in the present work are contained in the manuscript

