## supplementary material for "Synthesized Economic Evidence on the Cost-effectiveness of Screening Familial Hypercholesterolemia: A meta-analytic review and Aggregate Distributional Cost-Effectiveness Analysis" (from another version of this preprint: version 1, 2023-12-10)

### Supplementary materials

Appendix 1. Systematic Review and Data Analysis

*National and Analytical Perspective*

This study conducted a comprehensive analysis of 19 literature pieces, encompassing diverse geographical focuses: 32% in the UK, 16% in Australia, 16% in the US, 5% in Argentina, 5% in Poland, 11% in Spain, and 16% in the Netherlands. Geographically, 63% concentrated in Europe, 16% in North America, 16% in Oceania, and 5% in South America, with no studies from Asia and Africa.

These countries approached economic evaluations from different perspectives. Specifically, 79% of the studies solely analyzed from a healthcare perspective, considering direct medical costs, including screening and treatment expenses. In contrast, 5% of the studies conducted analyses from a societal perspective, while an additional 16% simultaneously considered both healthcare and societal perspectives. These studies not only factored in medical costs but also took into account indirect costs arising from productivity loss.

*Cost unit and discount*

The currency units used are US dollar, Euro, Australian dollar and British pound. 32% reported pounds, 26% euros, 16% Australian dollars and 26% US dollars. 89% discount cost and benefits to reflect loss of value in future outcomes, and only 11% do not discount. In the discounted papers, 82% of the cost and benefits adopted the same discount rate, and 18% adopted different discount values respectively. The cost discount rate ranges from 3%-6%, and the benefits discount rate ranges from 1%-5%.

*Economic Evaluation Methods, Outcome Measures, and Sensitivity Analysis*

In the study, 32% of the literature used cost-effectiveness analysis (CEA) as measured by the number of adverse events avoided and years of life gained (LYG), while 21% used cost-utility analysis (CUA) as measured by quality-adjusted life years (QALY) for health outcomes. Additionally, 47% combined CEA and CUA for economic evaluation. 68% reported QALY, 63% reported LYG, and 32% reported avoided adverse events.21% reported QALY, LYG, and CE simultaneously.

To ensure model robustness, 95% of studies conducted sensitivity analyses, with 42% using one-way sensitivity analyses and 53% utilizing both one-way and Probabilistic Sensitivity Analysis.

*Decision Analysis Models and Time horizon*

Various decision analysis modeling methods were employed, including Markov models, Decision trees, Life-table analysis, Simulated family trees, and combinations such as Decision tree + Markov model and Decision tree + Life-table analysis. Specifically, 16% used Markov models alone, 11% used Decision trees alone, 16% used Life-table analysis alone.

42% employed a combination of Decision tree + Markov model, and 11% combined

Decision tree + Life-table analysis.58% of studies utilizing Markov models reported Health States, with health state numbers ranging from 3-14.
Regarding the timeframe, 74% of studies focused on a lifetime, while some considered different tracking durations, including 10, 30, and 60 years, with 16% analyzing a 10-year span.

*FH Detection Methods, Sensitivity and Specificity, Treatment*
Different detection methods were employed, with 47% using gene testing, 5% using LDL-C (low-density lipoprotein cholesterol) testing, and 47% using both gene and LDL-C testing, demonstrating effectiveness.
During testing, 53% of studies reported gene testing sensitivity (ranging from 78.5% to 100%), and 47% reported specificity (ranging from 99.7% to 100%). In articles reporting both sensitivity and specificity, 32% assumed both to be 100%.
Post-screening treatment for FH patients commonly involved Statins (84%), followed by combinations like Statins and Ezetimibe (11%), and Statins, Ezetimibe, and PCSK9 (5%). Regarding adherence to lipid-lowering therapy, 21% of studies assumed 100% adherence.

Appendix 2. Type of health costs considered

| ID | Organization of screening | Consultation | Screening test | IC identification | Treatment | Follow up visits | Disease costs | Indirect costs |
|---|---|---|---|---|---|---|---|---|
| Kerr et al. (2017) | × | × | √ | √ | √ | × | √ | × |
| Crosland et al. (2018) | √ | √ | √ | √ | √ | × | √ | × |
| McKay et al. (2018) | √ | √ | √ | √ | √ | √ | √ | × |
| Marks et al. (2002) | √ | √ | √ | √ | √ | √ | √ | × |
| Marks et al. (2003) | √ | √ | √ | × | √ | × | √ | × |
| Nherera et al. (2011) | √ | √ | √ | × | √ | × | √ | × |
| Ademi et al. (2020) | √ | × | √ | √ | √ | √ | √ | × |
| Ademi et al. (2014) | × | × | √ | √ | √ | √ | √ | × |
| Marquina et al. (2022) | √ | × | √ | √ | √ | √ | √ | √ |
| Chen et al. (2015) | √ | √ | √ | √ | √ | √ | √ | √ |
| Spencer et al. (2022) | √ | × | √ | √ | √ | × | √ | × |

| | | | | | | | | |
|---|---|---|---|---|---|---|---|---|
| Jackson et al. (2021). | √ | √ | √ | × | √ | √ | × | × |
| Araujo et al. (2023) | × | √ | √ | √ | √ | √ | √ | × |
| Pelczarska et al.(2018) | √ | √ | √ | × | √ | √ | √ | × |
| Lázaro et al. (2017) | √ | × | √ | √ | √ | × | √ | √ |
| Oliva et al. (2009) | × | √ | √ | × | √ | √ | √ | × |
| Wonderling et al. (2004) | √ | √ | √ | √ | √ | √ | √ | × |
| Marang-van et al. (2002) | √ | √ | √ | √ | √ | √ | √ | × |
| Ademi et al. (2023) | √ | × | √ | √ | √ | √ | √ | √ |

√: Costs includes this.

×: This item is not included in the costs.

Appendix 3. CHEERS quality results

| CHEERS 2022 checklist | Item No | Kerr 2017 | Crosland,2018 | Mc Kay, 2018 | Marks,2002 | Marks,2003 | Nherera,2011 | Ademi,2020 | Ademi,2014 | Marquina, 2022 | Chen,2015 | Spencer,2022 | Jackson,2021 | Araujo,2023 | Pelczarska,2018 | Lázaro, 2017 | Oliva,2009 | Wonderling, 2004 | Marang-van 2002 | Ademi,2023 |
|---|---|---|---|---|---|---|---|---|---|---|---|---|---|---|---|---|---|---|---|---|
| Sum points | | 22.5 | 22 | 21 | 20 | 19 | 22 | 23.5 | 20.5 | 24.5 | 21 | 22.5 | 21.5 | 21 | 21 | 21 | 21.5 | 20 | 20.5 | 21.5 |
| **TITLE** | | | | | | | | | | | | | | | | | | | | |
| Title | 1 | 1 | 1 | 1 | 0.5 | 0.5 | 1 | 0.5 | 1 | 1 | 0.5 | 1 | 1 | 1 | 0.5 | 1 | 0.5 | 0.5 | 0.5 | 0.5 |
| **ABSTRACT** | | | | | | | | | | | | | | | | | | | | |
| Abstract | 2 | 1 | 1 | 1 | 1 | 1 | 1 | 1 | 1 | 1 | 1 | 1 | 1 | 1 | 1 | 1 | 1 | 0.5 | 1 | 1 |
| **INTRODUCTION** | | | | | | | | | | | | | | | | | | | | |
| Background and objectives | 3 | 1 | 1 | 1 | 1 | 1 | 1 | 1 | 1 | 1 | 1 | 1 | 1 | 1 | 1 | 1 | 1 | 1 | 1 | 1 |
| **METHODS** | | | | | | | | | | | | | | | | | | | | |
| Health economic analysis plan | 4 | 1 | 1 | 1 | 1 | 1 | 1 | 1 | 1 | 1 | 1 | 1 | 1 | 1 | 1 | 1 | 1 | 1 | 1 | 1 |
| Study population | 5 | 1 | 1 | 1 | 1 | 1 | 1 | 1 | 1 | 1 | 1 | 0.5 | 1 | 0.5 | 1 | 1 | 1 | 0.5 | 1 | 1 |
| Setting and location | 6 | 1 | 1 | 1 | 1 | 1 | 1 | 1 | 1 | 1 | 1 | 1 | 1 | 1 | 1 | 1 | 1 | 1 | 1 | 1 |
| Comparators | 7 | 1 | 1 | 1 | 1 | 1 | 1 | 1 | 1 | 1 | 1 | 1 | 1 | 1 | 1 | 1 | 1 | 1 | 1 | 1 |
| Perspective | 8 | 1 | 1 | 1 | 0.5 | 0.5 | 1 | 1 | 1 | 1 | 1 | 1 | 1 | 1 | 1 | 1 | 1 | 0.5 | 0.5 | 1 |
| Time horizon | 9 | 0.5 | 0.5 | 0.5 | 0.5 | 0.5 | 0.5 | 0.5 | 0.5 | 0.5 | 0.5 | 0.5 | 0.5 | 0.5 | 0.5 | 0.5 | 0.5 | 0.5 | 1 | 0.5 |
| Discount rate | 10 | 0.5 | 1 | 0.5 | 1 | 0 | 1 | 1 | 0.5 | 1 | 0.5 | 0.5 | 0.5 | 0.5 | 1 | 0.5 | 0.5 | 1 | 0.5 | 1 |

| | | | | | | | | | | | | | | | | | | | | |
|---|---|---|---|---|---|---|---|---|---|---|---|---|---|---|---|---|---|---|---|---|
| Selection of outcomes | 11 | 1 | 1 | 1 | 1 | 1 | 1 | 1 | 1 | 1 | 1 | 1 | 1 | 1 | 1 | 1 | 1 | 1 | 1 | 1 |
| Measurement of outcomes | 12 | 1 | 1 | 1 | 1 | 1 | 1 | 1 | 1 | 1 | 1 | 1 | 1 | 1 | 1 | 1 | 1 | 1 | 1 | 1 |
| Valuation of outcomes | 13 | 1 | 1 | 1 | 1 | 1 | 1 | 1 | 1 | 1 | 1 | 1 | 1 | 1 | 1 | 1 | 1 | 1 | 1 | 1 |
| Measurement and valuation of resources and costs | 14 | 1 | 1 | 1 | 1 | 1 | 1 | 1 | 1 | 1 | 1 | 1 | 1 | 1 | 1 | 1 | 1 | 1 | 1 | 1 |
| Currency, price date, and conversion | 15 | 1 | 1 | 1 | 0 | 0 | 1 | 1 | 1 | 1 | 1 | 1 | 1 | 0.5 | 0.5 | 1 | 1 | 1 | 1 | 1 |
| Rationale and description of model | 16 | 0.5 | 1 | 0.5 | 0.5 | 0.5 | 0.5 | 0.5 | 0.5 | 0.5 | 0.5 | 0.5 | 1 | 1 | 0.5 | 0.5 | 0.5 | 0.5 | 0.5 | 0.5 |
| Analytics and assumptions | 17 | 0.5 | 0.5 | 0.5 | 0 | 0.5 | 0.5 | 0.5 | 0.5 | 1 | 0.5 | 1 | 1 | 0 | 0.5 | 0.5 | 0.5 | 0.5 | 1 | 0.5 |
| Characterizing heterogeneity | 18 | 0 | 0 | 0 | 0 | 0 | 0 | 1 | 0 | 1 | 0 | 0 | 0 | 0 | 0 | 0 | 0 | 0 | 0 | 0 |
| Characterizing distributional effects | 19 | 1 | 0 | 0 | 1 | 1 | 0 | 1 | 0 | 1 | 0 | 1 | 1 | 1 | 0 | 0 | 1 | 1 | 1 | 1 |
| Characterizing uncertainty | 20 | 0.5 | 0.5 | 0.5 | 0.5 | 0.5 | 0.5 | 0.5 | 0.5 | 0.5 | 0.5 | 0.5 | 0 | 0.5 | 0.5 | 0.5 | 0.5 | 0.5 | 0.5 | 0.5 |

| | | | | | | | | | | | | | | | | | | | | |
|---|---|---|---|---|---|---|---|---|---|---|---|---|---|---|---|---|---|---|---|---|
| Approach to engagement with patients and others affected by the study | 21 | 0.5 | 0.5 | 0.5 | 0.5 | 0.5 | 0.5 | 0.5 | 0.5 | 0.5 | 0.5 | 0.5 | 0.5 | 0.5 | 0.5 | 0.5 | 0.5 | 0.5 | 0.5 | 0.5 |
| **RESULTS** | | | | | | | | | | | | | | | | | | | | |
| Study parameters | 22 | 1 | 1 | 1 | 1 | 1 | 1 | 1 | 1 | 1 | 1 | 1 | 0.5 | 1 | 1 | 1 | 1 | 1 | 1 | 1 |
| Summary of main results | 23 | 1 | 0.5 | 0.5 | 0.5 | 1 | 1 | 0.5 | 1 | 1 | 1 | 1 | 0.5 | 0.5 | 1 | 1 | 1 | 1 | 1 | 1 |
| Effect of uncertainty | 24 | 0.5 | 0.5 | 0.5 | 0.5 | 0.5 | 0.5 | 1 | 0.5 | 0.5 | 0.5 | 0.5 | 0 | 0.5 | 0.5 | 1 | 1 | 0.5 | 0.5 | 0.5 |
| Effect of engagement with patients and others affected by the study | 25 | 0 | 0 | 0 | 0 | 0 | 0 | 0 | 0 | 0 | 0 | 0 | 0 | 0 | 0 | 0 | 0 | 0 | 0 | 0 |
| **DISCUSSION** | | | | | | | | | | | | | | | | | | | | |
| Study findings, limitations, generalizability, and current knowledge | 26 | 1 | 1 | 1 | 1 | 1 | 1 | 1 | 1 | 1 | 1 | 1 | 1 | 1 | 1 | 1 | 1 | 1 | 1 | 1 |

| | | | | | | | | | | | | | | | | | | | | |
|---|---|---|---|---|---|---|---|---|---|---|---|---|---|---|---|---|---|---|---|---|
| Source of funding | 27 | 1 | 1 | 1 | 1 | 1 | 1 | 1 | 1 | 1 | 1 | 1 | 1 | 1 | 1 | 1 | 1 | 1 | 0 | 0 |
| Conflicts of interest | 28 | 1 | 1 | 1 | 1 | 0 | 1 | 1 | 0 | 1 | 1 | 1 | 1 | 1 | 1 | 1 | 0 | 0 | 0 | 1 |

Green: Yes

Light green: Partially reported

Red: Not reported

### Appendix 4. QHES instrument

| No. | Points | Kerr 2017 | Crosland,2018 | McKay,2018 | Marks,2002 | Marks,2003 | Nherera ,2011 | Ademi,2020 | Ademi,2014 | Marquina, 2022 | Chen, 2015 | Spencer,2022 | Jackson,2021 | Araujo,2023 | Pelczarska,2018 | Lázaro,2017 | Oliva,2009 | Wonderling,2004 | Marang-van 2002 | Ademi,2023 |
|---|---|---|---|---|---|---|---|---|---|---|---|---|---|---|---|---|---|---|---|---|
| 1 | 7 | 7 | 7 | 7 | 7 | 7 | 7 | 7 | 7 | 7 | 7 | 7 | 7 | 7 | 7 | 7 | 7 | 7 | 7 | 7 |
| 2 | 4 | 4 | 4 | 4 | 2 | 2 | 4 | 4 | 4 | 4 | 4 | 4 | 4 | 4 | 4 | 4 | 4 | 2 | 2 | 4 |
| 3 | 8 | 7 | 7 | 7 | 7 | 6 | 6 | 7 | 6 | 7 | 6 | 6 | 6 | 6 | 7 | 7 | 6 | 6 | 7 | 7 |
| 4 | 1 | 0 | 0 | 0 | 0 | 0 | 0 | 1 | 0 | 1 | 0 | 0 | 0 | 0 | 0 | 0 | 0 | 0 | 0 | 0 |
| 5 | 9 | 7 | 9 | 9 | 7 | 5 | 7 | 7 | 7 | 7 | 7 | 7 | 1 | 7 | 7 | 7 | 9 | 5 | 6 | 9 |
| 6 | 6 | 6 | 3 | 3 | 3 | 6 | 6 | 6 | 6 | 6 | 6 | 6 | 3 | 3 | 6 | 6 | 6 | 6 | 3 | 6 |
| 7 | 5 | 5 | 5 | 5 | 5 | 5 | 5 | 5 | 5 | 5 | 5 | 5 | 5 | 5 | 5 | 5 | 5 | 5 | 5 | 5 |
| 8 | 7 | 5 | 7 | 5 | 7 | 4 | 7 | 7 | 5 | 7 | 5 | 5 | 5 | 5 | 7 | 5 | 5 | 7 | 5 | 7 |
| 9 | 8 | 8 | 8 | 8 | 6 | 6 | 8 | 8 | 8 | 8 | 8 | 8 | 8 | 7 | 7 | 8 | 8 | 8 | 8 | 8 |
| 10 | 6 | 6 | 6 | 6 | 6 | 6 | 6 | 6 | 6 | 6 | 6 | 6 | 6 | 6 | 6 | 6 | 6 | 6 | 6 | 6 |
| 11 | 7 | 7 | 7 | 7 | 7 | 7 | 7 | 7 | 7 | 7 | 7 | 7 | 7 | 7 | 7 | 7 | 7 | 7 | 7 | 7 |
| 12 | 8 | 7 | 8 | 7 | 7 | 7 | 7 | 7 | 7 | 7 | 7 | 7 | 8 | 8 | 7 | 7 | 7 | 7 | 7 | 7 |
| 13 | 7 | 3 | 5 | 3 | 4 | 4 | 4 | 4 | 5 | 4 | 5 | 5 | 4 | 5 | 4 | 4 | 4 | 4 | 5 | 4 |
| 14 | 6 | 4 | 4 | 4 | 3 | 4 | 4 | 4 | 4 | 6 | 4 | 6 | 6 | 3 | 4 | 4 | 4 | 4 | 6 | 4 |
| 15 | 8 | 8 | 8 | 8 | 8 | 8 | 8 | 8 | 8 | 8 | 8 | 8 | 8 | 8 | 8 | 8 | 8 | 8 | 8 | 8 |
| 16 | 3 | 3 | 3 | 3 | 3 | 3 | 3 | 3 | 3 | 3 | 3 | 3 | 3 | 3 | 3 | 3 | 3 | 3 | 0 | 0 |

| | | | | | | | | | | | | | | | | | | | | |
|---|---|---|---|---|---|---|---|---|---|---|---|---|---|---|---|---|---|---|---|---|
| SUM | 100 | 87 | 91 | 86 | 82 | 80 | 89 | 91 | 88 | 93 | 88 | 90 | 81 | 84 | 89 | 88 | 89 | 85 | 82 | 89 |

75-100: high quality;

50-74： moderate quality;

25-49: low quality;

0-24: very low quality;

Appendix 5. Meta-analysis Data Extraction

Data were extracted according to different measure outcomes in the data synthesis study, and all currencies were converted into 2023 US dollars for calculation.

Cascade screening (2023 US dollar)

| Author Year | Incremental cost | QALY per gain | LYG per person | Total advents averted | Adverse events averted | Deaths averted |
|---|---|---|---|---|---|---|
| Kerr et al. (2017) | 4702.79 | 0.48 | NA | 46 MIs,50 cases of angina,8 strokes and 16 deaths | 104 | 16 |
| Marks et al. (2003) | 101486932.5 | NA | NA | 560 deaths | | 560 |
| Ademi et al. (2020) | -852.31 | 1.07 | 0.97 | 24.2 fewer acute non-fatal events and 7.55 death | 24.2 | 7.55 |
| Ademi et al. (2014) | 84620.28 | 29.07 | 24.95 | NA | NA | NA |
| Lázaro et al. (2017) | 39591894.47 | 767 | NA | 847Cardiac events;203Coronary deaths | 847 | 203 |
| Oliva et al. (2009) | 8922.69 | NA | 1.34 | NA | NA | NA |
| Wonderling et al. (2004) | 11433.06 | NA | 0.9 | NA | NA | NA |
| Ademi et al. (2023) | 31369.18 | 2.53 | 2.28 | NA | NA | NA |

Universal screening (2023 US dollar)

| Author Year | Incremental cost | QALY per gain | LYG per person | Total advents averted | Adverse events averted | Deaths averted |
|---|---|---|---|---|---|---|
| Marks et al. (2003) | 13500753.09 | NA | NA | 11.7 deaths | | 11.7 |
| Marquina et al. (2022) | 1061544599.77 | 51790 | 33488 | 3093 CHD events | 1813 | 1279 |
| Spencer et al. (2022) | 20836899.42 | 97.5 | 61.5 | NA | NA | NA |

Green: Reported QALYs;

Yellow: Reported LYGs;

Pink: Total CE;

Grey: Reported adverse events averted;

Blue: Reported deaths averted

Appendix 6. Comparative efficiency research (COMER)

$$ICER = \frac{c_1 - c_2}{e_1 - e_2} \quad (1)$$

where $1$ and $2$ correspond to the n new screening and reference screening. $c$ is cost. $e$ is effect. $ICER$ is Incremental cost-effectiveness ratio.

$$\begin{aligned} \text{TNHB} &= \sum \omega_s * NHB_s \\ &= \sum \omega_s * \left(k * (e_{s1} - e_{s2}) - (c_{s1} - c_{s2})\right) \\ &= k * \sum \omega_s * (e_{s1} - e_{s2}) - \sum \omega_s * (c_{s1} - c_{s2}) \end{aligned} \quad (2)$$

Where $s$ is the number of the study and $1$ and $2$ the new screening and reference screening. $c$ is cost. e is effect. $k$ is willingness to pay. $w$ is weight. $NHB$ is net health benefit. $TNHB$ is total net health benefit.
NHB and weight[1], respectively:

$$NHB_s = k * (e_{s1} - e_{s2}) - (c_{s1} - c_{s2}) \quad (3)$$

$$\begin{aligned} \omega_s &= \frac{1/var(NHB_s)}{\sum_{i=1}^{S}(1/var(NHB_i))} \\ &= \frac{1/(k^2 * Var(e_{s1} - e_{s2}) + Var(c_{s1} - c_{s2}) - 2 * k * Covar(e_{s1} - e_{s2}, c_{s1} - c_{s2})}{\sum_{i=1}^{S}(1/var(NHB_i))} \end{aligned} \quad (4)$$

Where $w$ is weight. $Var$ is variance. $k$ is willingness to pay.

Appendix 7. Population and disease distributional inputs

| | IMD1 (most deprived) | IMD2 | IMD3 | IMD4 | IMD5 (least deprived) | Source |
|---|---|---|---|---|---|---|
| Baseline distribution of health (QALE) | 63.21 | 67.61 | 69.95 | 73.10 | 75.00 | Love-Koh et al[2]. |
| Health opportunity costs distribution | 26% | 22% | 22% | 16% | 14% | Love-Koh et al[3]. |

The IMD, or Index of Multiple Deprivation, is a composite deprivation index used to measure health inequality. IMD1 refers to the most deprived areas, while IMD5 refers to the least deprived areas. Love-Koh et al.'s study estimated the baseline distribution of health (QALE) and the distribution of health opportunity costs. We utilized their data to calculate the Net Health Benefit of FH screening and the post-screening population distribution.
